# Isofurans and isoprostanes as markers of delayed brain injury and mitochondrial dysfunction following aneurysmal subarachnoid hemorrhage. A prospective observational study

**DOI:** 10.1101/2021.03.10.21252935

**Authors:** Joao A. Gomes, Ginger Milne, Asha Kallianpur, Leah Shriver

**Author notes:** Corresponding author*: Joao A. Gomes, MD, 9500 Euclid Ave., mail code S-80, Cleveland, OH 44195, Patients enrolled at Cleveland Clinic; Cleveland, OH. Biological samples analyzed at Vanderbilt University; Nashville, TN.

## Abstract

**Introduction:** F2-Isoprostanes (F2-IsoPs) and Isofurans (IsoF), specific markers of lipid peroxidation in vivo, have been reported to be elevated and have prognostic implications following subarachnoid hemorrhage (SAH). Platelet activation and vasoconstriction are attributed to these compounds. Elevated IsoF: F2-IsoPs ratios have been previously suggested as indicative of mitochondrial dysfunction. In this small study we examined their performance as specific biomarkers for delayed brain injury (DBI) development following SAH. We also explored if evidence of mitochondrial dysfunction could be found in a cohort of SAH patients.

**Methods:** Eighteen patients with SAH and 7 controls with normal neuroimaging and CSF analysis results underwent CSF sampling and abstraction of clinical, demographic, and laboratory data. Samples (two) of CSF were collected on day 1 and once on days 5-8 post-bleed. F2-IsoP and IsoF assays were performed at Vanderbilt Eicosanoid Core Lab by gas chromatography/mass spectroscopy. Levels are expressed in median (IQR) for non-parametric data. Repeated sample measurement were compared using the Wilcoxon signed-rank test, whereas the Mann Whitney test was used for other non-parametric data.

**Results:** Mean age was 61.2 ± 15.7 (SAH cases) vs. 47.6 ± 10 (controls) years, and 80% of SAH patients were female. Median Hunt-Hess score was 3 (2-4) and modified Fisher scale 3 (3-4). Thirty nine percent of patients developed DBI. F2-IsoP were significantly higher in SAH cases than in controls [47.5 (30.2-53.5) vs. 26.0 (21.2-34.5) pg/mL]. No significant differences were observed in patients with or without DBI [41 (33.5-52) vs. 44 (28.5-55.5) pg/mL]. IsoF were elevated in the second CSF sample in 9 patients, but undetectable in the remainder cases and all controls. Patients who developed DBI had significantly higher IsoF than cases who did not [(57 (34-72) vs. 0 (0-34) pg/mL]. Patients who met criteria for delayed injury had a significantly higher IsoF: F2IsoPs ratio on the late CSF sample [1.03 (1-1.38) vs. 0 (0-0.52)].

**Conclusions:** Preliminary findings from this study suggest that IsoF may represent a specific biomarker predicting DBI following SAH and provide possible evidence of CNS mitochondrial dysfunction in SAH. Future studies to further explore the value of IsoF as biomarkers of secondary brain injury and the contribution of mitochondrial dysfunction and ferroptosis to clinical outcomes following SAH seem warranted.

## Introduction

Occurrence of delayed brain injury (DBI) following SAH is traditionally assessed by means of regular clinical examinations which carries limitations due to poor patient condition in some instances, examiner subjectivity, and overall low sensitivity [1]. Transcranial Doppler (TCD), while commonly used as a screening tool for the detection of vasospasm, has at best moderate specificity and predictive value [2]. In light of these challenges, there is an urgent need for the development of suitable biomarkers for DBI prediction. Although a panel of genetic and non-genetic markers showed initial promise at predicting DBI occurrence, prospective validation is still lacking [3].

The non-enzymatic oxidation of polyunsaturated fatty acids such as arachidonic acid leads to the formation of isoprostanes (F_2_-IsoPs) and isofurans (IsoF). Isofurans formation is favored over F_2_-IsoPs under increasing oxygen concentrations. Collectively, these compounds have emerged as one of the most reliable markers to assess oxidative stress in vivo, and given their procoagulant and vasospastic properties may potentially act as mediators of oxidative injury following SAH [4]. While previous studies have examined levels of F2-IsoPs and IsoF and their relationship to oxidative stress following SAH [5, 6, 7], their use as specific biomarkers for the development of DBI has not been explored to date.

The ratio of IsoF to F2-IsoPs has been proposed as a specific marker of mitochondrial dysfunction [8], and given the presence of increased redox-active iron in the central nervous system (CNS) following SAH [9], the possibility of intracellular iron accumulation, mitochondrial injury and triggering of ferroptosis, we wanted to explore if evidence of mitochondrial dysfunction and DBI could be found in a cohort of SAH patients.

## Methods

Patients admitted to the neurointensive care unit with the diagnosis of aneurysmal SAH were screened for enrollment in an observational prospective cohort study seeking to assess oxidative stress in this population. Subjects were approached for participation in the study if they were between 18 and 80 years of age, admission Hunt and Hess grade was 2-5, had a clear time of onset of symptoms, and required insertion of an external ventricular drain (EVD) as part of their clinical management. High grade patients not expected to survive the index admission were not approached for participation.

Screening for DBI and clinical management were dictated by the clinical team and in accordance with current management guidelines [10]. Samples of cerebrospinal fluid (CSF) were obtained from the EVD within the first 24 hours (early sample), and once between days 5 and 8 (late sample) following the onset of ictus. Development of DBI was defined as a focal neurologic impairment (i.e. hemiparesis, neglect, apraxia, aphasia, or hemianopia) or unexplained reduction in Glasgow Coma Scale of > 2 points lasting at least one hour, not apparent immediately following aneurysm occlusion and not attributable to other conditions (i.e. hydrocephalus, seizures, infections, etc.). Demographic, laboratory, and clinical data from subjects enrolled in this study were abstracted for the duration of the hospitalization. Control CSF was obtained from patients with suspected neurological disease and seen at the “lumbar puncture clinic” that resulted normal after testing analysis and imaging studies. This study was conducted in accordance with all local IRB guidelines, and informed consent was obtained from all individual participants included in the study.

Levels of F_2_-IsoPs and IsoF in the CSF were measured at the Vanderbilt Eicosanoid Core Lab by gas chromatography/mass spectroscopy as previously described [11]. Levels are expressed in median (IQR) for non-parametric data, and as means ± SD if they followed a normal distribution. T-test was used to compare independent means, whereas Chi-square for categorical variables. Repeated sample measurements were compared using the Wilcoxon signed-rank test, whereas the Mann-Whitney test was used to compare continuous, non-normally distributed data.

## Results

A total of 18 SAH patients and 6 controls are included in this preliminary report. No sample size calculations were feasible since no prior data exist to help guide such analysis. This represent our first effort to further refine next steps. The sample size was primarily driven by time and monetary constraints. Baseline demographics, disease severity at presentation, clinical course, and outcomes are outlined in *table 1*. Control subjects were overall younger than SAH patients, and included fewer women. Early CSF samples could only be obtained in 9/18 patients (50%), while late samples were available for the whole cohort. Criteria for DBI were met by 7/18 (39%). A combination of hemodynamic augmentation, intra-arterial vasodilator therapy, and angioplasty were used to treat DBI in this cohort. A slight majority of patients were discharged home or to an acute rehab, while 2 patients died during the first 30 days.

**Table 1.** Demographic and clinical characteristics. AR: acute rehab; DBI: delayed brain injury; EVD: external ventricular drain; GCS: Glasgow coma scale; HH: Hunt and Hess grade; LOS: length of stay; mFS: modified Fisher scale; mRS: modified Rankin scale

|  | SAH (n=18) | Controls (n=6) |
| --- | --- | --- |
| Age (years) | 61.2 ± 15.7 | 47.6 ± 10 |
| Female (%) | 78 | 67 |
| Smoking (%) | 60 | 33 |
| HH | 3 (2-4) |  |
| mFS | 3 (3-4) |  |
| GCS | 12.5 (9-14) |  |
| DBI | 7 (39%) |  |
| - Median time to DBI (days) | 6.5 (2-7) |  |
| - Hemodynamic augmentation | 6/7 |  |
| - IA vasodilators | 5/7 |  |
| - Angioplasty | 2/7 |  |
| ICU LOS (days) | 12.5 (10-18) |  |
| Hospital LOS (days) | 13.5 (10-18) |  |
| Duration of EVD (days) | 16 (14-20) |  |
| 30-day mortality | 2 (11%) |  |
| Discharged home or to AR (%) | 56 |  |
| 90-day mRS | 2 (1-4) |  |

Routine CSF analysis in the control group yielded the following results: protein 43 (37-44) mg/dL, glucose 62 (57-67) mg/dL, WBC 1 (1-2) cells/mm^3^, and RBC 0.5 (0-1) cells/mm^3^. The most common indications for a lumbar puncture were to rule out demyelinating disease and as part of the work up for peripheral neuropathy. In all control individuals, CNS pathology was ultimately deemed not to be present by the primary neurologist. A non-significant elevation of CSF F_2_-IsoP levels over time in the SAH cohort was observed [34 (30.5-42.5) vs. 48 (28.5-53.0) pg/mL], and at the end of the study period CSF F2-IsoPs were significantly higher compared to controls [47.5 (30.2-53.5) vs. 26.0 (21.2-34.5) pg/mL, *p <*0.05; *table 2*]. No significant differences were observed in patients with or without DBI [41 (33.5-52) vs. 44 (28.5-55.5) pg/mL]. Isofurans were not detected in control samples, and only in one instance in the early samples in a patient who eventually met criteria for DBI. Nine SAH patients had elevated IsoF in the late CSF sample and subjects who developed DBI had significantly higher IsoF levels than those who did not [(57 (34-72) vs. 0 (0-34) pg/mL, *p*<0.05]. The presence of CSF IsoF had 86% sensitivity to detect DBI, with a negative predictive value of 89%.

**Table 2.** Levels of Isoprostanes (F_2_-IsoPs) and isofurans (IsoF) in cerebrospinal fluid (CSF) at early (24 hours) and late (5-8 days) time points following ictus, as well as controls. Shaded areas indicative of development of delayed brain injury.

| Subjects | Early CSF<br>F<sub>2</sub>IsoPs<br>(pg/mL) | Late CSF<br>F<sub>2</sub>IsoPs<br>(pg/mL) | Early CSF<br>IsoF<br>(pg/mL) | Late CSF<br>IsoF<br>(pg/mL) | Late<br>Isof:F<sub>2</sub>IsoPs<br>ratio (late) |
| --- | --- | --- | --- | --- | --- |
| SAH 1 | - | 67 | - | 72 | 1.07 |
| SAH 2 | - | 61 | - | 0 | 0 |
| SAH 3 | - | 59 | - | 57 | 0.96 |
| SAH 4 | - | 49 | - | 0 | 0 |
| SAH 5 | 33 | 28 | 0 | 0 | 0 |
| SAH 6 | 44 | 29 | 0 | 0 | 0 |
| SAH 7 | 22 | 65 | 0 | 34 | 0.52 |
| SAH 8 | - | 44 | - | 45 | 1 |
| SAH 9 | - | 47 | - | 36 | 0.76 |
| SAH 10 | - | 34 | - | 34 | 1 |
| SAH11 | 34 | 50 | 0 | 38 | 0.76 |
| SAH 12 | 28 | 28 | 0 | 0 | 0 |
| SAH 13 | 61 | 54 | 0 | 0 | 0 |
| SAH 14 | 34 | 39 | 0 | 0 | 0 |
| SAH 15 | 41 | 52 | 42 | 72 | 1.38 |
| SAH 16 | 36 | 29 | 0 | 72 | 2.48 |
| SAH 17 | - | 48 | - | 0 | 0 |
| SAH 18 | - | 26 | - | 0 | 0 |
| Control 1 | 21 | - | 0 | - | 0 |
| Control 2 | 22 | - | 0 | - | 0 |
| Control 3 | 39 | - | 0 | - | 0 |
| Control 4 | 36 | - | 0 | - | 0 |
| Control 5 | 21 | - | 0 | - | 0 |
| Control 6 | 30 | - | 0 | - | 0 |

Patients who met criteria for delayed injury had a significantly higher IsoF: F_2_IsoPs ratio on the late CSF sample [1.03 (1-1.38) vs. 0 (0-0.52), *p*<0.05]. Moreover, IsoF: F_2_IsoPs ratios of ≥; 0.9 were associated with 100% specificity and positive predictive value for the detection of DBI. Since exposure to higher oxygen concentrations can favor the production of IsoF over F_2_IsoPs, we also abstracted results of arterial blood gas analyses that were performed as part of routine clinical care leading up to the late CSF sample collection date. The median arterial oxygen tension (PaO_2_) was 115 (77.5-176) mmHg in the non-DBI group, and 130.5 (89.2-189.7) mmHg in the DBI cohort, a difference which failed to achieve statistical significance (*p*=0.23).

## Discussion

The main results of this exploratory investigation is that CSF IsoF, and in particular the IsoF:F_2_- IsoPs ratio may serve as a specific marker of secondary brain injury following SAH. Furthermore, the significantly higher ratio of IsoF:F_2_-IsoPs in DBI cases may represent evidence of mitochondrial dysfunction in these patients.

The pathology of SAH is complicated by early brain insults related to the initial aneurysm rupture and subsequent damage associated with secondary processes such as delayed injury. The pathophysiology of DBI has not been fully elucidated, but vasospasm, inflammation, cortical spreading depolarization, iron-induced oxidative stress, and thrombosis of the microvasculature among others have been implicated [9, 12].

Serial clinical examination and TCD are the traditional ways of assessing for DBI, but both carry significant limitations. Ultrasound is operator dependent, only screens for one of the putative causes of DBI, and has at best moderate predictive value [2]. Although a panel of biomarkers has been proposed, there is limited evidence backing their use, up to 21 different molecules were included, and they lack prospective validation [3]. The development of a relatively simple and highly specific biomarker that can be assessed serially would be a welcome addition to both the clinical care and research endeavors surrounding SAH patients.

The free radical-mediated formation of prostaglandin–like compounds has emerged as one of the most reliable approaches to assessing oxidative stress status in vivo [4]. Besides their role as markers of endogenous oxidative injury, F_2_-IsoPs have also been shown to have potent biological actions, including cerebral vasoconstriction and platelet activation through thromboxane receptors [14]. When oxidative injury takes place in the setting of elevated oxygen tension, a carbon-centered radical is attacked by oxygen giving rise to IsoF instead of F_2_-IsoP [4]. In vivo, mitochondrial dysfunction is the other known setting in which high oxygen tension is encountered (due to decreased oxygen consumption). Therefore, an increased IsoF:F_2_-IsoPs ratio has been postulated as a specific marker of mitochondrial injury and oxidative stress [8, 13].

Previously, small studies had already identified elevation of F_2_-IsoP in serum, urine, and CSF following SAH and attempted correlations with clinical outcomes [5, 6]. In a pilot study, CSF F_2_-IsoP were positively correlated with poor outcome or severity of clinical condition [6]. In another small investigation, urinary levels of F_2_-IsoPs were associated with development of vasospasm and worse neurologic outcomes [5]. In contrast, we did not find any significant correlation between CSF F_2_-IsoPs and clinical outcomes of aneurysmal SAH. This might at least in part be explained by different sampling timeline as well as the use of different clinical endpoints, such as DBI.

The presence of IsoF in the CSF of SAH patients has been shown in a small clinical study [7], however no potential association with clinical endpoints was reported. Our preliminary investigation suggests that IsoF may be a rather specific marker of secondary brain injury after SAH. Moreover, for the first time evidence for the possibility of mitochondrial dysfunction (i.e. increased CSF IsoF: F_2_IsoPs ratio) following SAH is presented. Given the increased availability of redox-active iron in the CNS after SAH [9] and the progressive accumulation of nonheme iron in brain tissue after aneurysm rupture [15], it is likely that the local conditions are present for iron-induced mitochondrial dysfunction and/or the triggering of ferroptosis. While this particular mechanism is beyond the scope of the present investigation, this intriguing hypothesis represents a new line of inquiry that is worth pursuing in the future.

It is important to acknowledge several limitations of the present study. First, the small sample size limits the generalizability of the preliminary findings, and prospective clinical validation is warranted before these biomarkers can be adopted as part of routine clinical care. For the same reason, better estimates of positive and negative predictive value and sensitivity for prediction of DBI after aneurysmal SAH require significantly larger sample N. Second, the control group was not truly asymptomatic. Obvious ethical reasons made it difficult to obtain CSF from healthy individuals, and therefore, we did our best to rule out CNS involvement in the control group. Also, it should be pointed out that there is a sex imbalance between SAH cases and controls. Whether sex affects levels of CSF IsoF or F2-IsoPs in the setting of acute SAH is currently unknown.

Third, in half of SAH patients, an early CSF sample could not be obtained, usually due to the initial absence of an EVD, disease severity requiring extended neuroimaging, time in the operating room or angiography suite, or lack of availability of study personnel during non-work hours. Fourth, There are potential confounders that we were unable to adjust for in our analysis (i.e. age, sex, race/ethnicity, smoking status, substance use, etc.) given our small sample size. Lastly, our endpoint was based on a clinical definition for DBI. The lack of a clear endpoint when conducting SAH research is an issue that plagues the field, and whether special neuroimaging (i.e., restricted diffusion lesions on MRI) represents a better predictive marker for DBI needs further investigation.

## Conclusion

Preliminary findings from this study suggest that IsoF may represent a specific biomarker predicting DBI following SAH and provide possible evidence of CNS mitochondrial dysfunction in SAH. Future studies to further explore the value of IsoF as biomarkers of secondary brain injury and the contribution of mitochondrial dysfunction and ferroptosis to clinical outcomes following SAH seem warranted.

## Data Availability

All data presented and discussed in the present manuscript is available and currently kept in a password-protected, HIPPA compliant, secured database in a hospital server.

This study was funded by a grant from The Aneurysm and AVM Foundation (TAAF). The authors declare that they have no conflicts of interest.

